# Research on the Diagnostic Value and Immune Microenvironment Regulatory Mechanism of FOLR3 Gene in Endometrial Cancer Based on Multi-omics Data Algorithms

**DOI:** 10.64898/2025.12.09.25341245

**Authors:** Xu Xianyun, Wang Jiaxi, Zhang Hongyu, Wang Yunyun, Gao Xue lin, Xin Wenhu, Qin Tiansheng

## Abstract

**Background and Objective:** FOLR3 serves as an important member of the folate metabolic pathway and plays a crucial role in various malignant tumors. However, the expression pattern, diagnostic value, and regulatory mechanism of FOLR3 in endometrial cancer (EC) remain unclear. This study aimed to explore the expression characteristics, clinical significance, and related molecular regulatory networks of FOLR3 in EC through bioinformatics analysis.

**Methods:** TCGA datasets served as training sets, while GSE17025 from GEO served as validation sets. FOLR3 differential expression was analyzed using Wilcoxon rank-sum test, diagnostic efficacy evaluated by ROC curves, and prognostic value assessed via Kaplan-Meier survival analysis. Candidate genes were identified through WGCNA, univariate Cox regression, and differential expression analysis. Key genes were screened using machine learning algorithms (RF, LASSO, SVM-RFE) and PPI network analysis. ceRNA regulatory networks were constructed, and immune infiltration was analyzed using CIBERSORT.

**Results:** FOLR3 was significantly overexpressed in EC (*P* < 0.01) with diagnostic AUC values > 0.6 in both datasets. High FOLR3 expression indicated poor prognosis (HR=2.5, *P* < 0.05). Among 5,539 differentially expressed genes, 3 key genes (AURKA, POLQ, CDKN2A) were identified via multi-algorithm screening. Enrichment analysis showed involvement in cell division, p53 signaling, and cellular senescence. The ceRNA network comprised 72 nodes and 169 relationships, with KCNQ1OT1 and XIST as key lncRNA regulators. FOLR3 positively correlated with memory B cells and M0 macrophages, negatively with naive B cells and resting mast cells, with significant immune score differences (*P* < 0.05).

**Conclusion:** FOLR3 was significantly upregulated in EC, serving as an effective diagnostic biomarker and independent prognostic predictor. FOLR3 participated in tumorigenesis via complex ceRNA networks and regulated the tumor immune microenvironment. This study provides novel molecular targets and theoretical foundation for precision diagnosis and therapy of EC.

## 1 Introduction

Endometrial cancer (EC) is one of the most common gynecological malignancies. With increasing life expectancy and improved living standards, adverse dietary habits and obesity have contributed to a rising incidence of EC, posing a significant threat to women’s health. EC is among the few malignancies with both rising incidence and mortality rates[1]. Over the past decade, the incidence has shown a gradual upward trend, and in some regions of China as well as certain Western countries, EC has become the most common gynecological malignancy[1–3]. Although most EC patients have a relatively favorable prognosis, patients with advanced or recurrent disease face poor outcomes[3]. Early-stage EC is predominantly managed with surgical resection; however, chemotherapy, radiotherapy, and hormonal therapy show limited efficacy in advanced, recurrent, or metastatic EC[4, 5]. Therefore, the identification of highly specific and sensitive diagnostic biomarkers and prognostic predictive factors for chemotherapy, immunotherapy, and targeted therapy is crucial for achieving early diagnosis and individualized precision treatment of EC.

In recent years, precision medicine based on molecular classification has played an increasingly important role in EC treatment. In 2013, The Cancer Genome Atlas (TCGA) research network first proposed a molecular classification system for EC[6]. Subsequently, ProMisE and TransPORTEC consortia developed and validated immunohistochemistry-based molecular surrogate classifiers, providing practical and cost-effective molecular diagnostic approaches for clinical application[7, 8]. However, existing stratification systems are primarily based on limited molecular markers and clinicopathological parameters, with restricted accuracy, necessitating the discovery of additional clinically valuable biomarkers[9].

Folate Receptor 3 (FOLR3) is a member of the specialized folate receptor family and participates in intracellular folate transport and one-carbon metabolism together with FOLR1 and FOLR2[10]. Unlike FOLR1/FOLR2, FOLR3 lacks glycosylphosphatidylinositol (GPI) anchor modification and exists as a secretory protein in the circulatory system[11, 12], with predominant expression in hematopoietic tissues[11, 13]. With advancing research, FOLR3 has been found to be closely associated with the development of malignant tumors, serving as a diagnostic and radiotherapy sensitivity biomarker for non-small cell lung cancer[14–16]. Furthermore, Yuan et al. demonstrated that FOLR1 and FOLR3 expression levels are significantly higher in ovarian cancer compared to breast cancer and malignant mesothelioma, indicating that differential expression of folate receptor genes possesses important diagnostic value[17]. Emerging evidence suggests that folate receptors not only participate in one-carbon metabolism but also directly regulate multiple signaling pathways, including JAK-STAT3 and ERK1/2, playing important roles in tumorigenesis and development[18]. However, the expression characteristics, clinical significance, and mechanistic roles of FOLR3 in EC remain unclear.

With the rapid development of high-throughput sequencing technologies and bioinformatics methods, cancer research based on multi-omics data has become a current research hotspot. Machine learning algorithms have demonstrated tremendous potential in cancer biomarker discovery and prognostic prediction, enabling the identification of key features from complex high-dimensional data[19, 20]. Competing endogenous RNAs (ceRNAs) participate in tumorigenesis and development through lncRNA-miRNA-mRNA regulatory axes and play important roles in various cancers, providing new perspectives for understanding the regulatory mechanisms of tumorigenesis[21, 22]. In recent years, multiple studies have successfully applied machine learning algorithms to construct cancer prognostic models based on ceRNA networks, providing novel strategies for precision tumor therapy[23, 24].

The tumor immune microenvironment plays a critical role in cancer development and therapeutic response. Immunotherapy has emerged as a breakthrough in cancer treatment; however, only a subset of patients benefit from these approaches[25]. A comprehensive understanding of the characteristics and regulatory mechanisms of the tumor immune microenvironment is essential for improving immunotherapy efficacy. The development of algorithms such as CIBERSORT has made it possible to infer immune cell infiltration in tumor tissues through transcriptomic data, providing powerful tools for investigating the tumor immune microenvironment[26]. Recent studies have demonstrated the superior performance of network biology and machine learning-based approaches in predicting immunotherapy response, offering new strategies for developing individualized immunotherapy[27].

Based on the aforementioned background, this study aimed to systematically investigate the expression characteristics, diagnostic value, and prognostic significance of FOLR3 in EC by integrating multi-omics data from TCGA and Gene Expression Omnibus (GEO) databases and employing multiple machine learning algorithms and bioinformatics methods. Furthermore, by constructing ceRNA regulatory networks and analyzing the immune microenvironment, we sought to elucidate the molecular regulatory mechanisms of FOLR3, providing new molecular targets and theoretical basis for precision diagnosis and treatment of EC, which has important implications for advancing individualized therapy of EC.

## 2 Methods

### 2.1 Data Download and Preprocessing

Endometrial cancer (EC)-related datasets were downloaded from the TCGA database (https://www.cancer.gov/ccg/research/genome-sequencing/tcga) as training sets, which contained 24 normal control samples and 177 EC patient tumor samples. The EC-related transcriptome dataset GSE17025 was downloaded from the GEO database (https://www.ncbi.nlm.nih.gov/geoprofiles/) as validation sets. The GSE17025 dataset was based on the GPL570 platform and included 91 EC and 12 normal tissue samples. EC genome-wide association study (GWAS) data were sourced from the GWAS database (https://storage.googleapis.com/ukb-b-13545.vcf), which included 461,782 control samples and 1,151 EC patient samples. With genes as exposure factors, GWAS datasets were obtained from the OpenGWAS database (https://gwas.mrcieu.ac.uk/).

### 2.2 Transcriptome Data Differential Expression Analysis

Based on the training set, the limma package was used with thresholds of |log2 fold change| > 2 and *P* < 0.05 to perform differential analysis and screen differentially expressed genes (DEGs), and volcano plots and heatmaps were generated.

### 2.3 Univariate Cox Analysis and Mendelian Randomization Analysis

Univariate Cox regression analysis was conducted to assess the association between gene expression levels and EC prognosis. Mendelian randomization (MR) analysis using the TwoSampleMR package identified EC risk-associated genes, with expression quantitative trait loci (eQTLs) as instrumental variables. Causal effects were estimated using inverse variance weighted (IVW) and MR-Egger methods. Genes with P < 0.05 were considered significantly associated with EC risk.

### 2.4 FOLR3 Gene Differential Analysis and Kaplan-Meier Survival Curve Analysis

The Wilcoxon rank-sum test was used to compare FOLR3 expression between EC and CON groups. Receiver operating characteristic (ROC) curves evaluated diagnostic efficacy (AUC > 0.6 indicating good discrimination). Kaplan-Meier survival analysis assessed the prognostic value of FOLR3 expression. P < 0.05 was considered statistically significant.

### 2.5 Clinical Factor Analysis

Clinical factors (age, clinical stage, overall survival) were extracted from TCGA and analyzed descriptively. Chi-square test or Fisher’s exact test compared categorical variables; t-test or Mann-Whitney U test compared continuous variables; Spearman correlation was used for ordinal variables. Multiple comparison correction was applied with P < 0.05 as significance threshold.

### 2.6 Weighted Gene Co-expression Network Analysis (WGCNA)

WGCNA was performed to identify co-expressed gene modules. The optimal soft threshold β was determined using the pickSoftThreshold function. Gene modules were constructed with parameters: minimum module size = 30, deep split = 3, maximum module distance = 0.25. Pearson correlation identified modules significantly associated with EC (P < 0.05). Hub genes were selected with |GS| > 0.5 and |MM| > 0.5. The intersection of DEGs, hub genes, and Cox-identified genes yielded candidate genes.

### 2.7 Candidate Gene Enrichment Analysis

Gene Ontology (GO) and Kyoto Encyclopedia of Genes and Genomes (KEGG) pathway enrichment analysis were performed using the DAVID database (P < 0.05 for significance). Gene set enrichment analysis (GSEA) identified signaling pathways differentially expressed between EC and normal groups.

### 2.8 Screening of Key Genes Related to FOLR3 Through PPI Algorithm and Machine Learning Algorithms

Four algorithms were applied to screen candidate genes: three machine learning algorithms [Random Forest (RF), Least Absolute Shrinkage and Selection Operator (LASSO), and Support Vector Machine Recursive Feature Elimination (SVM-RFE)] and one network algorithm [Maximum Clique Centrality (MCC)]. The intersection of genes identified by all four algorithms was considered key genes.

### 2.9 Protein-Protein Interaction (PPI) Network Analysis and FOLR3 Gene ceRNA Network Construction

PPI networks were constructed using the STRING database and visualized in Cytoscape. For ceRNA network construction, miRNAs targeting key genes were predicted using the miRWalk database, and upstream lncRNAs were identified using starBase. The lncRNA-miRNA-mRNA network was visualized in Cytoscape to reveal regulatory mechanisms.

### 2.10 Immune Infiltration Analysis

The CIBERSORTx algorithm estimated relative abundance of 22 immune cell subtypes in EC and control samples. Wilcoxon rank-sum test compared immune cell proportions between groups. Pearson correlation analyzed correlations between immune cells and FOLR3 expression. ESTIMATE algorithm calculated tumor microenvironment scores (stromal score, immune score, overall score). Violin plots compared these scores between high and low FOLR3 expression groups.

### 2.11 Statistical Analysis

R programming language (v 4.2.2) was used for bioinformatics analysis. Wilcoxon rank-sum test was employed for comparison of differences between two groups. *P* < 0.05 was considered statistically significant.

## 3 Results

### 3.1 Transcriptome Level Differential Expression Analysis

Compared with the CON group, a total of 5,539 genes were significantly differentially expressed in the EC group, including 2,291 upregulated genes and 3,248 downregulated genes (Figure 1A). The heatmap showed the expression of the top 15 upregulated and downregulated genes (Figure 1B).

**Figure 1.**
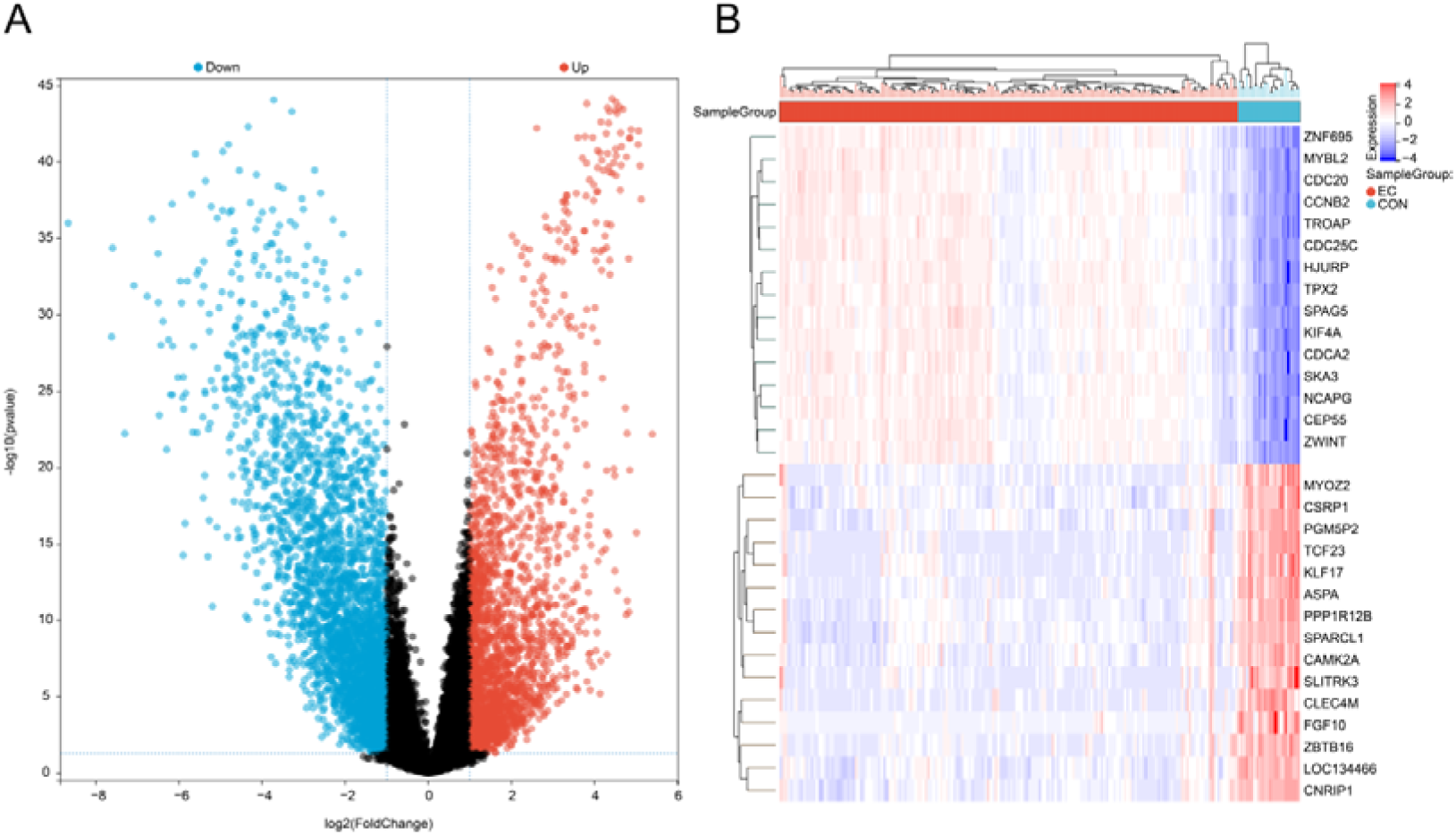
Transcriptome Data Expression Differences Between Endometrial Cancer (EC) and Control (CON) Groups. (A) Volcano plot; (B) Expression heatmap of top 15 upregulated and downregulated differential genes.

### 3.2 Univariate Cox and Mendelian Randomization Analysis

Through univariate Cox analysis and differentially expressed gene analysis, 177 candidate genes were obtained (Supplementary Figure 1). Further intersection analysis using Mendelian randomization methods with relevant genes (Supplementary Figure 2) yielded four intersecting genes: EZR, SLCO4C1, FOLR3, and ARHGEF3. Since the FOLR3 gene has not been previously reported in the literature, this study selected FOLR3 as the research target to further explore the molecular regulatory mechanism between this gene and EC.

### 3.3 FOLR3 Expression Validation and Kaplan-Meier Survival Curve Analysis

Based on TCGA transcriptome data and GEO dataset GSE17025, the Wilcoxon rank-sum test was used to analyze the expression of FOLR3 in EC and CON groups, and the diagnostic efficacy of FOLR3 for EC was explored through ROC analysis. Expression analysis results showed that FOLR3 expression was significantly upregulated in the EC group compared to the CON group in both TCGA and GEO datasets (*P* < 0.01) (Figure 2A, 2B). ROC analysis results showed that the AUC values of FOLR3 for distinguishing EC and CON groups were greater than 0.6 in both TCGA and GEO datasets (Figure 2C, 2D). These results indicated that FOLR3 might play an important role in the occurrence and development of EC and had potential diagnostic value. Meanwhile, the association between FOLR3 gene expression and prognostic value in EC was investigated. The relationship between EC survival outcomes and FOLR3 gene expression was evaluated through Kaplan-Meier cumulative curves from the TCGA database. Results showed that the survival rate of the high-expression group was significantly lower than that of the low-expression group (Figure 4E), with a hazard ratio HR = 2.5, *P* < 0.05, indicating that patients in the high-expression group had a higher risk of death than those in the low-expression group, and the survival difference between the two groups was statistically significant. These findings indicated that the FOLR3 gene could serve as an effective independent marker for predicting EC prognosis.

**Figure 2.**
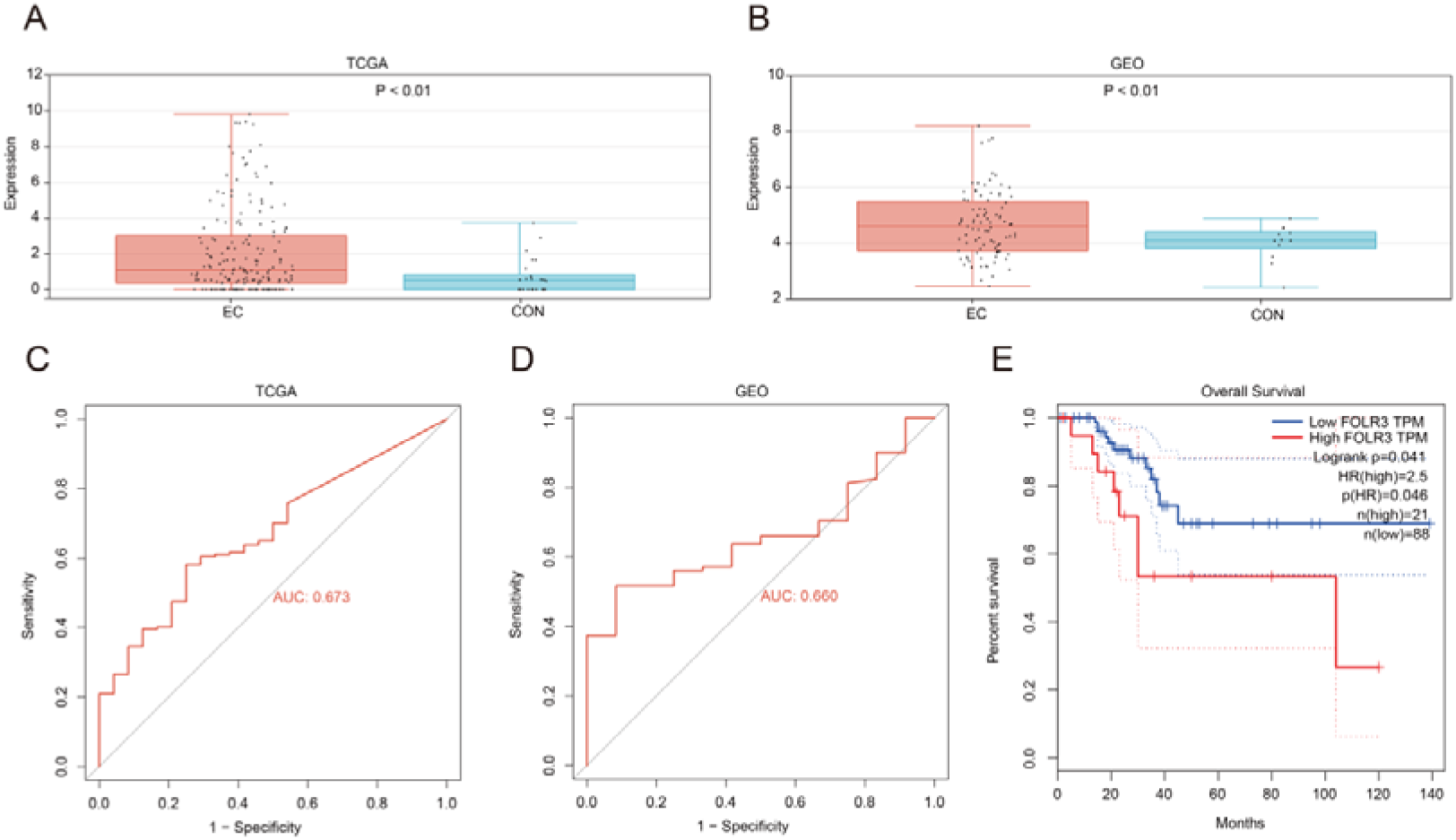
FOLR3 Gene Differential Analysis and Kaplan-Meier Survival Curve Analysis. (A) FOLR3 expression in the TCGA database. (B) FOLR3 expression in the GEO database. (C) ROC curve analysis of FOLR3 in the TCGA database. (D) ROC curve analysis of FOLR3 in the GEO database. (E) Kaplan-Meier survival curve analysis of FOLR3.

**Figure 3.**
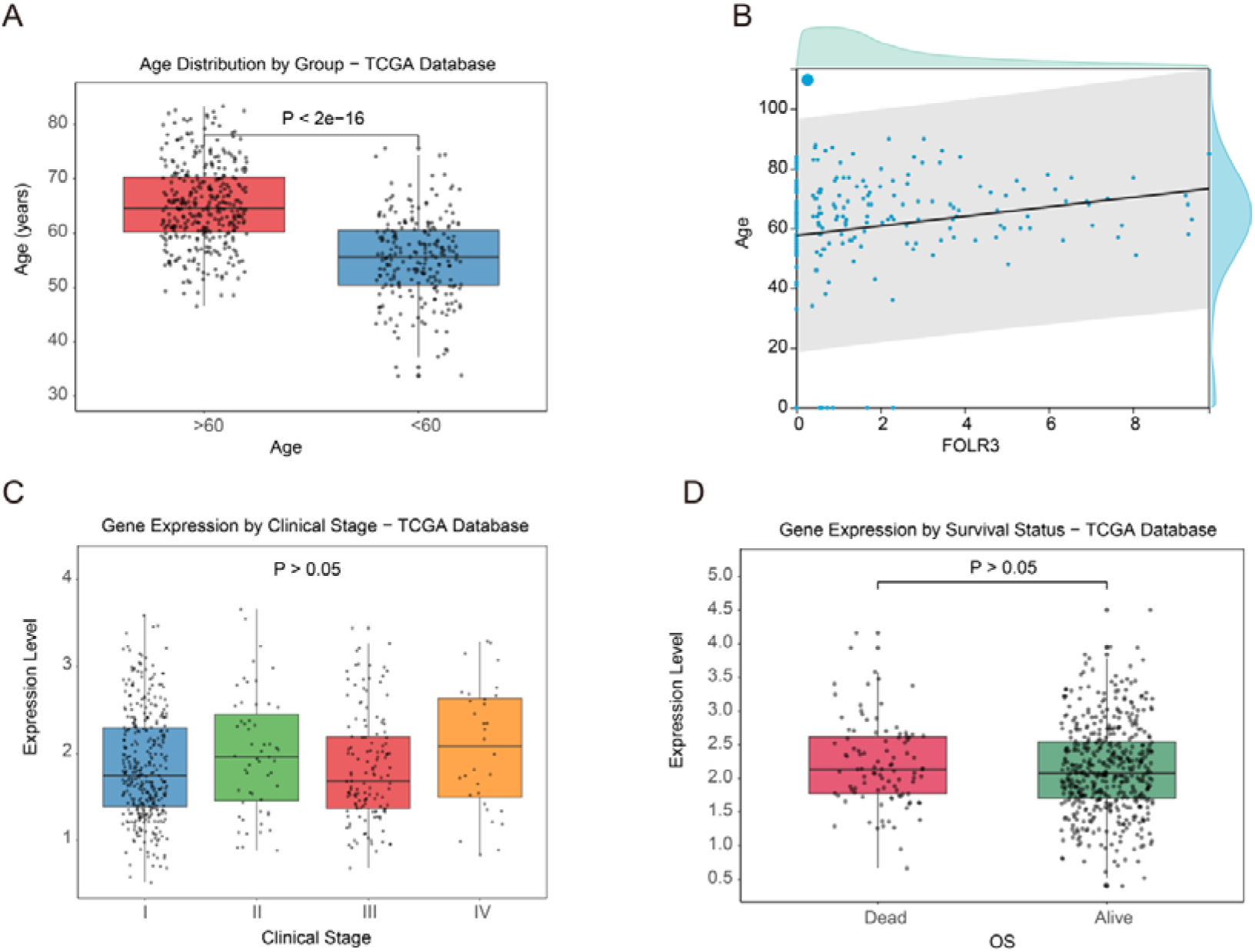
Box Plots of Clinical Factor Analysis. (A) Comparison of expression levels across different age groups. (B) Scatter plot of the correlation between Age and FOLR3.(C) Expression level comparison between different clinical stage. (D) Tumor level comparison among patients with different OS overall survival periods.

**Figure 4.**
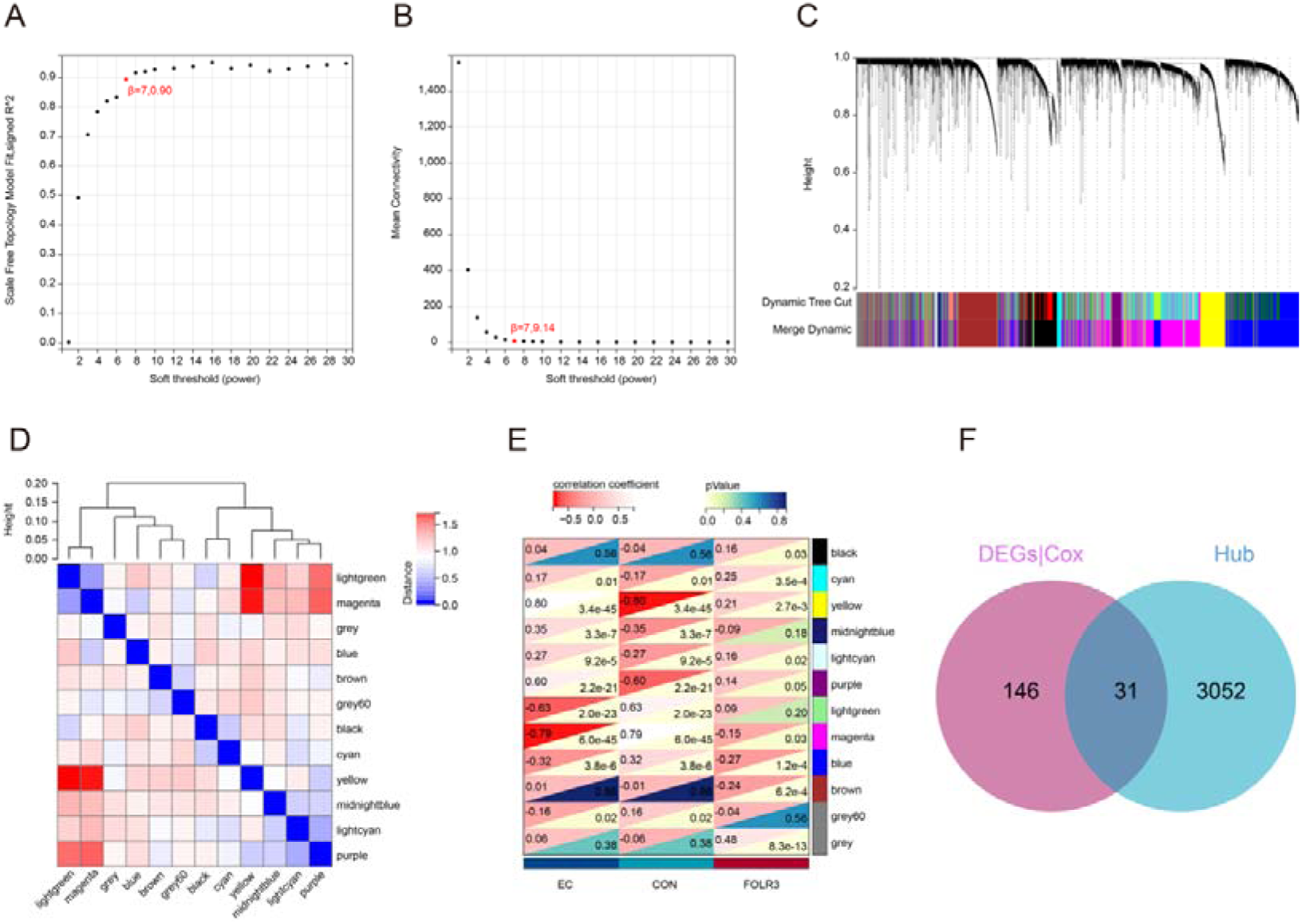
WGCNA. (A) Trend of Scale Free Topology Model Fit R² with soft threshold β; (B) Trend of Mean Connectivity with β; (C) Gene module clustering diagram; (D) Module eigenvector clustering diagram; (E) Module-phenotype correlation heatmap; (F) Venn diagram of candidate genes and Hub genes.

### 3.4 Clinical Factor Analysis

Based on the analysis of the TCGA database, the correlation between FOLR3 expression and clinicopathological parameters (Figure 5) revealed that the age distribution of patients in the >60-year-old group was significantly higher than that in the <60-year-old group (P < 0.05) (Figure 5A). The expression level of the FOLR3 gene showed a weak positive correlation trend with patient age, but the correlation was weak and the data points were scattered (Figure 5B). However, there was no significant difference in FOLR3 expression levels among patients at different clinical stages (Stage I, II, III, IV) (P > 0.05), with similar median expression levels and substantial overlap in the box plots across stages (Figure 5C). Similarly, there was no statistically significant difference in FOLR3 expression levels between the deceased (Dead) and alive (Alive) patient groups (P > 0.05) (Figure 5D). In summary, FOLR3 gene expression showed a weak correlation with age but no significant association with clinical stage or overall survival status. This suggests that FOLR3 may play a role early in the disease process or participate in pathological mechanisms independent of staging. Its value as an independent prognostic indicator requires further validation.

**Figure 5.**
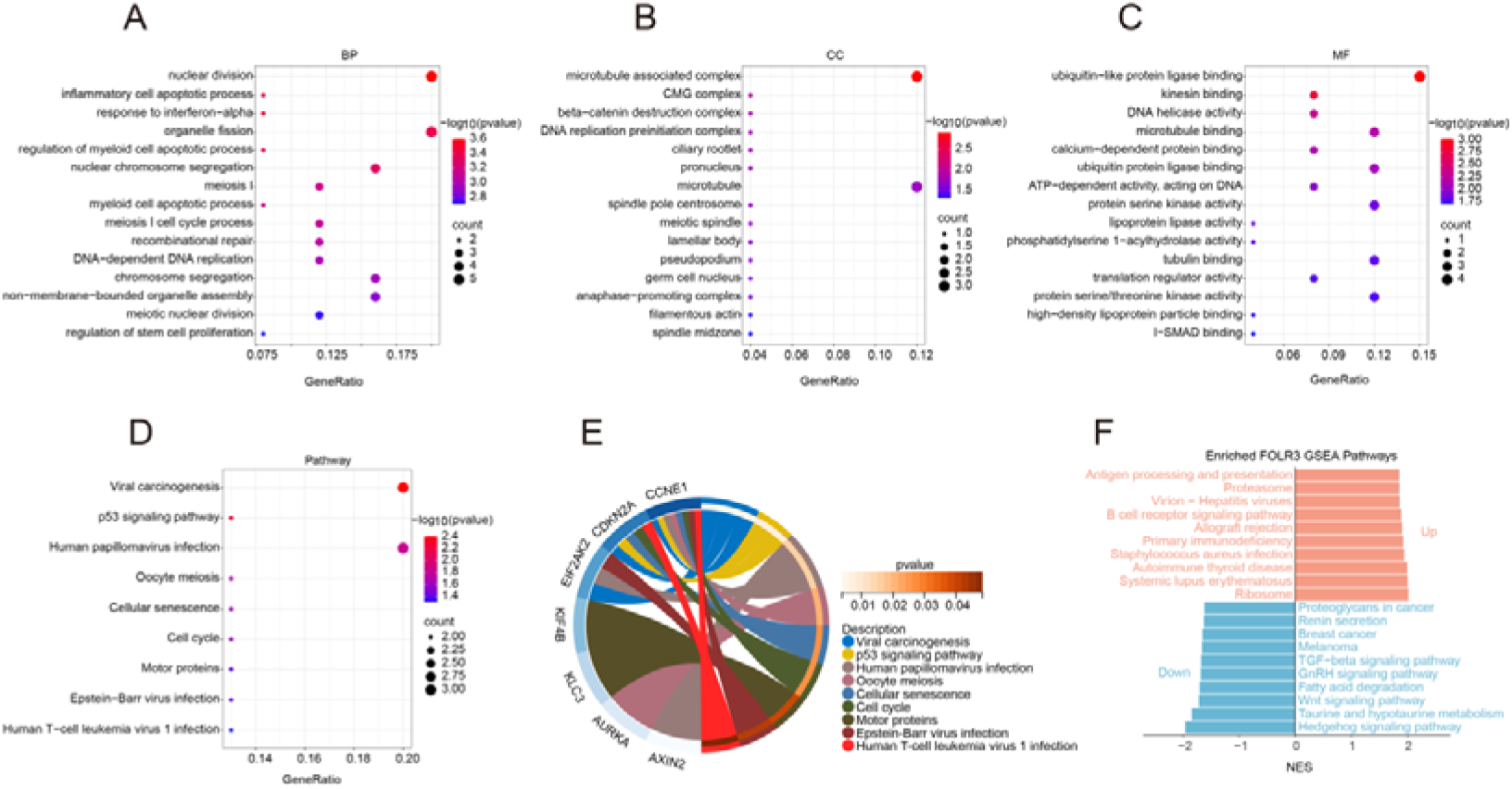
Functional Enrichment Analysis. (A-C) Bubble plots of BP (A), CC (B), MF (C) GO terms enriched by candidate genes (top 10 most significant terms). (D) Bubble plot of KEGG pathways enriched by candidate genes. (E) Connection relationships between co-expressed genes and enriched KEGG pathways. (F) Single-gene GSEA analysis of FOLR3.

### 3.5 WGCNA Analysis

An adjacency matrix was constructed based on the selected optimal soft threshold (β = 7) (Figure 4A, 4B) and further transformed into a topological overlap matrix (TOM) to reflect the co-expression similarity between genes. Then, based on hierarchical clustering and dynamic tree cutting algorithms, 12 gene modules were generated (Figure 4C), namely black, cyan, yellow, midnightblue, lightcyan, purple, lightgreen, magenta, blue, etc. Subsequently, the connectivity of module eigengenes (MEs) was analyzed, and results showed that the distance between modules was greater than 0.25, indicating good independence between each module (Figure 4D). Correlation analysis showed that 5 modules were significantly correlated with EC phenotype and FOLR3 (*P* < 0.05) (Figure 4E), and 3,083 Hub genes were extracted and identified from these gene modules. By comparing candidate genes and Hub genes and taking their intersection, 31 candidate genes were identified (Figure 6F), which were considered to be co-expressed genes related to FOLR3 in EC.

**Figure 6.**
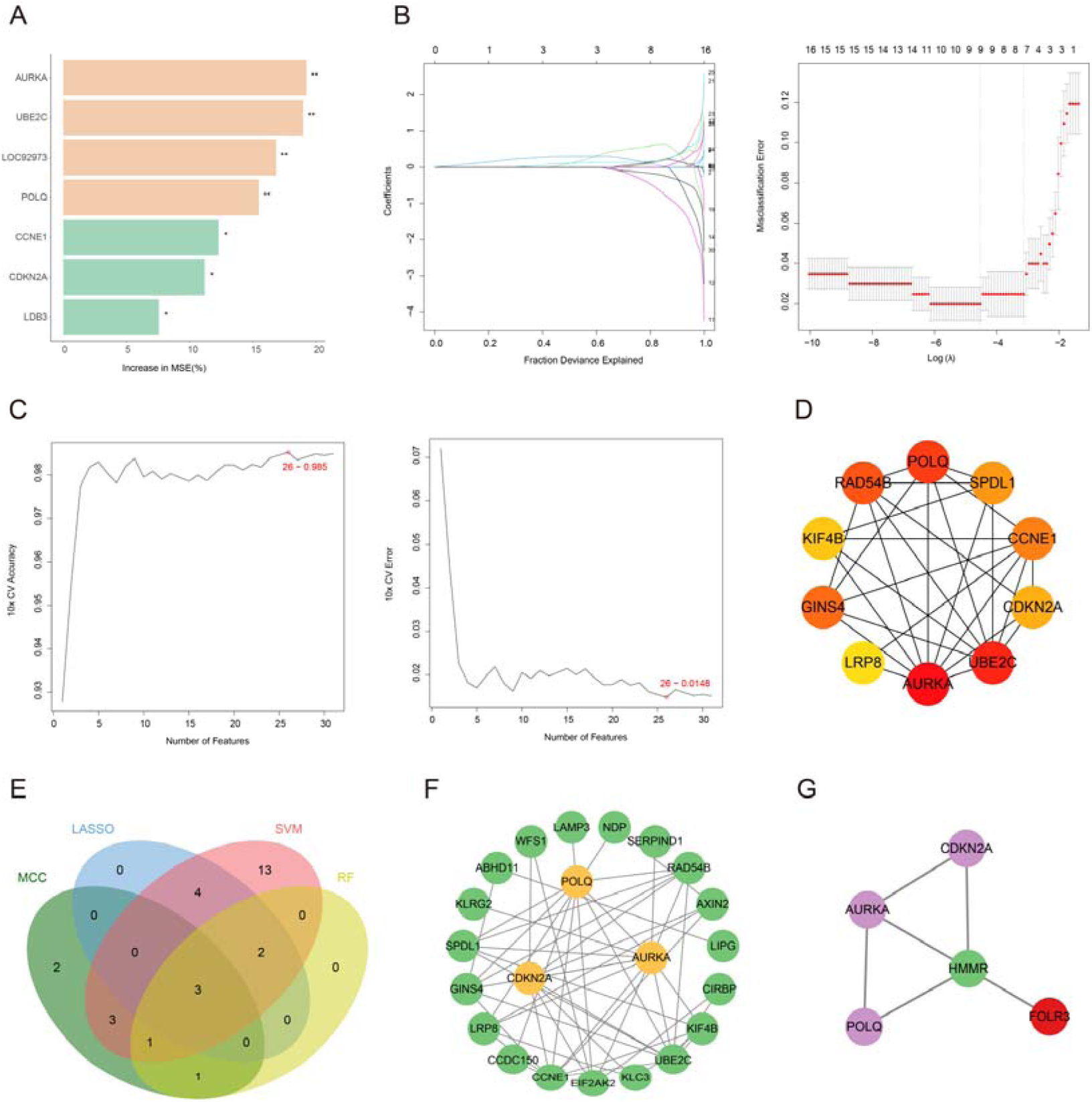
Key Gene Screening and Regulatory Network Relationships. (A-D) Screening results of candidate genes using Random Forest (A), LASSO (B), SVM-RFE (C), and PPI-MCC (D) algorithms. (E) Venn diagram showing the intersection of feature genes identified by the four algorithms. (F) PPI network, green nodes represent indirectly related genes. (G) Indirect regulatory relationships between FOLR3 (green) and the three key genes (purple).

### 3.6 Common Gene Enrichment Analysis

GO and KEGG functional enrichment analysis was performed on the 31 screened common genes. In GO enrichment analysis, these genes were mainly enriched in biological processes (BP) such as nuclear division, inflammatory cell apoptotic process, response to interferon-alpha, organelle fission, regulation of myeloid cell apoptotic process, and nuclear chromosome segregation (Figure 5A); cellular components (CC) such as microtubule associated complex, CMG complex, beta-catenin destruction complex, DNA replication preinitiation complex, ciliary rootlet, and pronucleus (Figure 5B); and molecular functions (MF) such as ubiquitin-like protein ligase binding, kinesin binding, DNA helicase activity, microtubule binding, calcium-dependent protein binding, and ubiquitin protein ligase binding (Figure 5C). KEGG enrichment analysis showed that these genes participated in signaling pathways including Viral carcinogenesis, p53 signaling pathway, Human papillomavirus infection, Oocyte meiosis, Cellular senescence, and Cell cycle (Figure 5D, 5E). Through single-gene GSEA analysis of FOLR3 (Figure 5F), the FOLR3 gene exhibited a complex multi-level regulatory network. FOLR3 had important associations with cellular behavior regulatory pathways such as cell adhesion and migration.

### 3.7 Key Gene Screening

Four machine learning algorithms (RF, LASSO, SVM) and PPI analysis were used to screen candidate genes (Figure 6A-D). In the training set, the Random Forest algorithm identified 7 feature genes, LASSO obtained 9 feature genes, SVM-RFE identified 26 feature genes, and PPI-MCC algorithm selected the top 10 genes. Intersection analysis of the four algorithms ultimately identified 3 key genes: AURKA, POLQ, and CDKN2A (Figure 6E). Additionally, STRING database analysis revealed that 31 common genes formed a PPI network containing 23 nodes and 49 edges (Figure 6F), suggesting these genes may be key regulatory genes involved in endometrial cancer progression. Further analysis demonstrated indirect regulatory relationships between the FOLR3 gene and the three key genes (Figure 6G).

### 3.8 Construction of Nomogram

Then, a nomogram risk prediction model was constructed based on these 3 key genes and the FOLR3 single gene, and it was found that high expression of the 4 key genes was a risk factor for EC (Figure 7A). The calibration curve showed good consistency between predicted probability and actual occurrence probability, indicating that the model had high predictive accuracy (Figure 7B). Additionally, Decision Curve Analysis (DCA) evaluated that the nomogram had good clinical benefit (Figure 7C).

**Figure 7.**
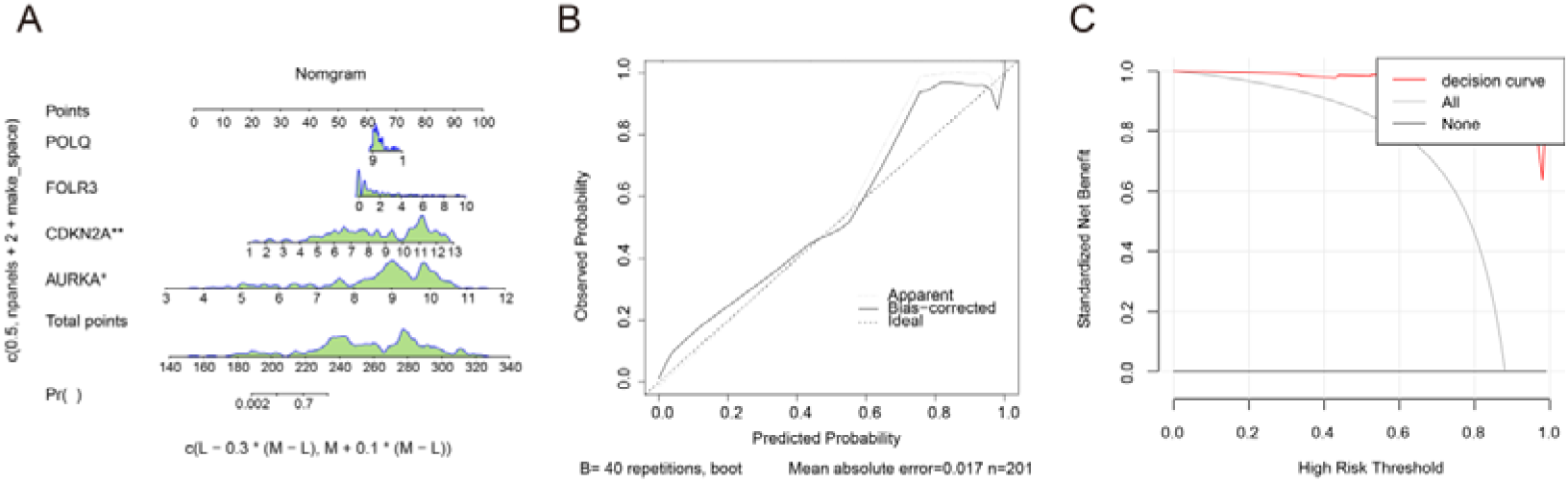
Nomogram and Decision Curves. (A) Nomogram model constructed with key genes. (B) Calibration curve evaluating model accuracy. (C) Model decision curve.

### 3.9 Construction of Regulatory Networks

The ceRNA network contained a total of 1 key mRNA, 6 miRNAs, 65 lncRNAs and 169 interaction relationships. Among them, KCNQ1OT1 and XIST were the lncRNAs with the largest degree values in the ceRNA network topology; hsa-miR-130a-3p and hsa-miR-130b-3p were the miRNAs with the largest degree values in the ceRNA network; FOLR3 was the mRNA with the largest degree value in the ceRNA network (Supplementary Figure 3).

### 3.10 FOLR3 Involvement in EC Immune Cell Analysis

The CIBERSORT method was used to analyze immune cell infiltration in EC and control group samples. A total of 22 immune cell types were identified (Figure 8A). Correlation analysis revealed the strongest positive correlation between Neutrophils and Activated Mast cells (r = 0.450, P < 0.05), and the strongest negative correlation between Resting NK cells and Activated NK cells (r = -0.496, P < 0.05) (Figure 8B). In EC samples, Follicular Helper T cells, Plasma cells, Activated Dendritic cells, and Regulatory T cells (Tregs) were significantly upregulated (Figure 8C), while M2 Macrophages, Naive B cells, Activated NK cells, and Resting Mast cells were significantly downregulated. Correlation analysis between FOLR3 gene expression and immune cell infiltration showed that FOLR3 was significantly positively correlated with Memory B cells and M0 Macrophages, and significantly negatively correlated with Naive B cells and Resting Mast cells (P < 0.05) (Figure 8D). Further analysis of FOLR3 expression distribution across different tumor microenvironment scoring categories (Figure 8E) demonstrated different distribution patterns between high and low FOLR3 expression groups in each scoring category, with significant differences in the ImmuneScore (P < 0.05).

**Figure 8.**
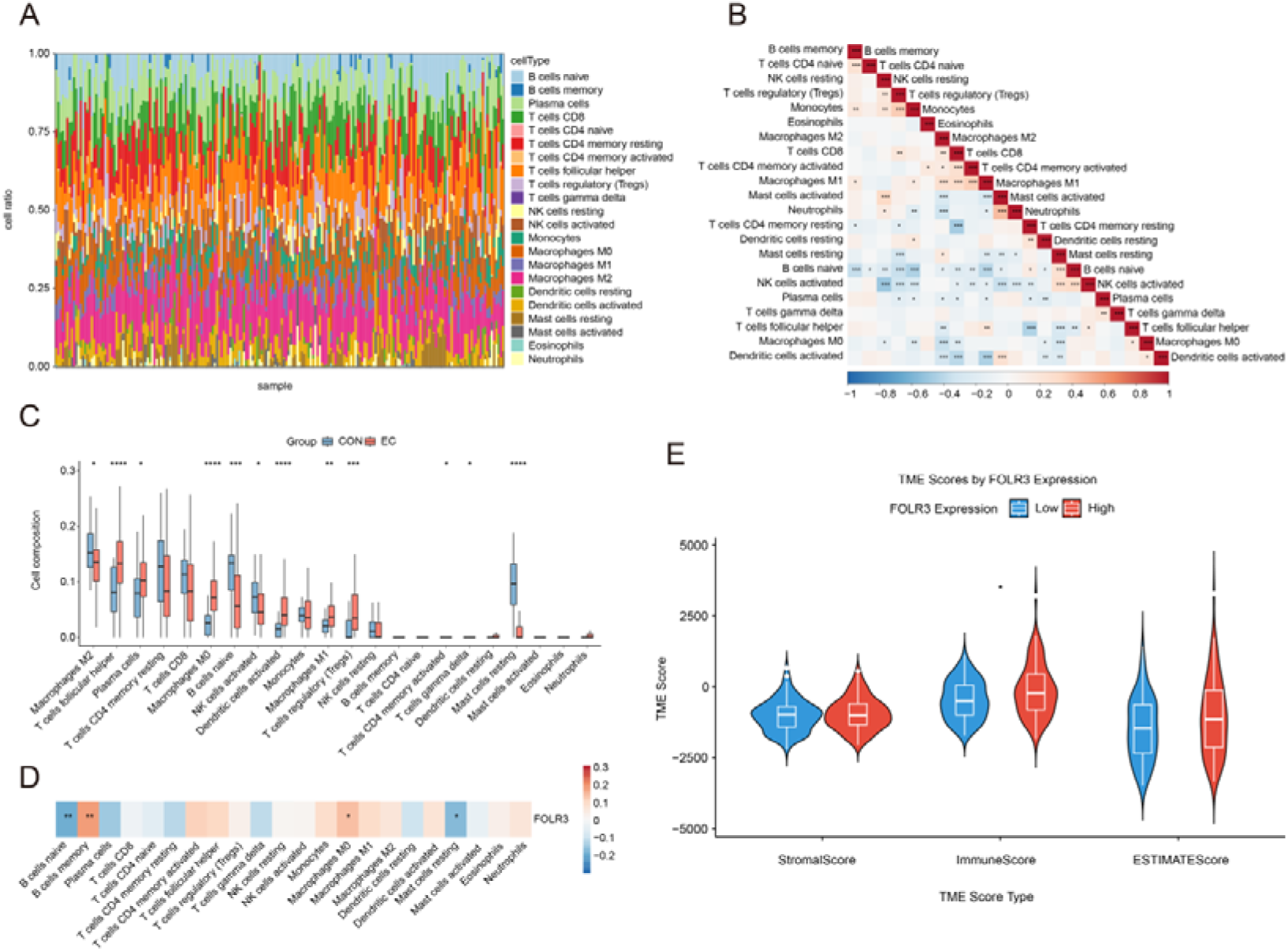
Immune Infiltration Analysis. (A) Immune cell infiltration proportions among samples. (B) Correlation analysis heatmap between different immune cells, \**P* < 0.05, \*\**P* < 0.01, \*\*\**P* < 0.001. (C) Differential analysis of immune cell infiltration proportions between EC and CON group samples \**P* < 0.05, \*\**P* < 0.01, \*\*\**P* < 0.001, \*\*\*\**P* < 0.0001. (D) Correlation heatmap between FOLR3 gene and immune cells, \**P* < 0.05, \*\**P* < 0.01. (E) Relationship analysis between FOLR3 gene expression levels and TIME scores, \**P* < 0.05.

## 4 Discussion

This study is the first to systematically elucidate the critical role of FOLR3 in EC. We found that FOLR3 is significantly upregulated in EC tissues. Additionally, the diagnostic AUC values exceeded 0.6 in two independent datasets, suggesting the potential of FOLR3 as an auxiliary diagnostic biomarker for EC. Kaplan-Meier survival analysis demonstrated that high FOLR3 expression is associated with poor patient prognosis, providing new molecular evidence for clinical risk stratification. FOLR3 expression showed correlation with age but no significant association with clinical stage or overall survival, suggesting that its prognostic value is primarily reflected in disease progression risk assessment. By integrating machine learning algorithms and the MCC algorithm in PPI networks, we successfully identified three key genes—AURKA, POLQ, and CDKN2A—that synergistically interact with FOLR3.

FOLR3, as a folate receptor, plays a critical role in tumor metabolism by mediating folate uptake. Folate is an important cofactor of one-carbon metabolism, providing carbon units for DNA and RNA synthesis and participating in nucleotide biosynthesis[12]. Folate participates in DNA methylation by providing methyl donors, and aberrant DNA methylation is closely associated with tumor suppressor gene silencing and oncogene activation[28, 29]. Recent studies suggest that FOLR3 may also participate in non-classical signal transduction. Upon folate binding, FOLR3 can activate pro-proliferative signaling pathways such as Src/focal adhesion kinase (FAK) and phosphatidylinositol 3-kinase/protein kinase B/mechanistic target of rapamycin (PI3K/AKT/mTOR), which have been demonstrated to be hyperactivated in EC progression[30]. This suggests that FOLR3 may function as a multifunctional molecule regulating tumor proliferation and migration. Therefore, FOLR3 plays a central role in EC development through dual mechanisms of metabolic reprogramming and signal transduction regulation.

Aurora kinase A (AURKA), a serine/threonine protein kinase family member, plays a pivotal regulatory role in cell division, particularly in spindle formation, centromere maturation, and cell cycle checkpoint control[31]. Extensive research has demonstrated that AURKA is significantly upregulated in EC tissues, with its overexpression closely associated with tumor grade, myometrial invasion depth, and lymph node metastasis, serving as an independent risk factor for poor prognosis[32–34]. Our PPI network analysis revealed a protein-protein interaction between AURKA and FOLR3, suggesting that these two proteins may synergistically regulate cell division to promote aberrant proliferation of EC cells. Furthermore, AURKA inhibitors may serve as prognostic predictors and biomarkers for EC treatment, opening new directions for targeted therapy[35].

DNA polymerase theta (POLQ) is a key enzyme involved in DNA damage repair. Its aberrant expression often leads to compromised DNA damage repair capacity, thereby inducing chromosomal instability[36–38]. POLQ is abnormally overexpressed in numerous malignancies and is closely associated with tumor invasiveness and chemotherapy resistance[39–41]. In EC, POLQ mutations are predominantly enriched in advanced tumors and promote chromosomal instability, malignant tumor progression, and treatment resistance through error-prone DNA repair mechanisms[42]. In our study, POLQ upregulation further confirmed its important role in EC. Similarly, POLQ inhibitors have demonstrated synthetic lethal effects in homologous recombination repair-deficient tumors[43], providing new strategies for precision treatment of EC.

Cyclin-dependent kinase inhibitor 2A (CDKN2A) is a key regulator of the G1/S checkpoint of the cell cycle[44]. In most cancers, CDKN2A functional loss (through mutation, deletion, or promoter methylation) is a common early event[45]. However, in EC, CDKN2A exhibits complex expression patterns and functional roles. Some studies report that high CDKN2A expression in EC is associated with favorable prognosis[46]. Interestingly, in our study, CDKN2A functioned as a risk gene (hazard ratio [HR] > 1), which may reflect that during EC progression, CDKN2A function has undergone abnormal changes, or its upregulation represents a stress response to DNA damage but is no longer able to effectively inhibit tumor progression. Therefore, the role of CDKN2A in EC requires comprehensive evaluation by integrating specific molecular backgrounds and clinicopathological characteristics.

Functional enrichment analysis further confirmed the synergistic mechanisms of these key genes. Although human papillomavirus (HPV) infection is primarily associated with cervical cancer, recent studies have revealed that HPV infection may also participate in some EC cases, particularly through inactivation of p53 and retinoblastoma (Rb) pathways by viral E6/E7 proteins, disrupting cell cycle regulation[47]. Inactivation of the p53 pathway is often accompanied by abnormal activation of multiple pro-proliferative signaling pathways such as PI3K/AKT/mTOR and mitogen-activated protein kinase (MAPK), forming complex signal network interactions that collectively drive malignant EC progression[48]. Notably, single-gene GSEA of FOLR3 revealed that FOLR3 is primarily involved in interactive regulation of cell behavior-related pathways such as cell adhesion and migration. Aberrant expression of cell adhesion molecules is a key initiating event in tumor invasion and metastasis. Through regulation of the epithelial-mesenchymal transition (EMT) process, tumor cells lose polarity and intercellular connections and acquire migratory and invasive capacity[49]. Activation of the cell migration pathway not only promotes local tumor infiltration and distant metastasis but is also closely associated with angiogenesis, matrix remodeling, and immune evasion[50]. This multi-pathway cross-talking network structure indicates that FOLR3 is not a single functional executor but rather a critical regulatory hub that integrates signal inputs from multiple biological processes including cell cycle, inflammatory immunity, and cell adhesion-migration, maintaining dynamic balance of cellular function through synergistic effects on multiple pathways.

KCNQ1OT1 and XIST, serving as the lncRNAs with the highest degree values, play central roles in the expression regulation of FOLR3. KCNQ1OT1 has been confirmed to exert oncogenic effects in multiple cancers[51]. XIST regulates the expression of tumor-associated genes through ceRNA mechanisms in various cancers[52, 53]. In EC, a female-specific malignancy, aberrant XIST expression may possess special biological significance, and its regulatory relationship with FOLR3 may reflect sex-specific molecular regulatory mechanisms. The miR-130 family has been demonstrated to exert either tumor-suppressive or pro-tumorigenic effects in multiple cancers[54], and changes in its expression levels may directly impact the mRNA stability and translation efficiency of FOLR3. Thus, the construction of ceRNA networks provides a novel perspective for understanding tumor heterogeneity[55, 56].

Different patients may harbor differential ceRNA networks, and such differences may be an important factor contributing to the variable prognosis among EC patients with identical histological types. In the future, precision treatment strategies based on individualized ceRNA network characteristics may become an important component of personalized tumor medicine.

Immune cell infiltration analysis based on the CIBERSORT algorithm revealed the critical role of FOLR3 in regulating the tumor immune microenvironment of EC. Correlation analysis showed that FOLR3 expression was significantly positively correlated with memory B cells and M0 macrophages, while significantly negatively correlated with naive B cells and resting mast cells. The enrichment of M0 macrophages merits particular attention. Macrophages exhibit classical M1/M2 polarization patterns, with M1 macrophages possessing pro-inflammatory and anti-tumor properties, while M2 macrophages displaying anti-inflammatory and pro-tumor characteristics; M0 macrophages represent unpolarized macrophages[57]. The positive correlation between FOLR3 and M0 macrophages suggests that in high FOLR3-expressing EC, macrophages may exist in a “neutral” or “undecided” functional state, unable to effectively exert anti-tumor effects yet not fully transitioning to the pro-tumor M2 phenotype. This “undecided” immune functional state may provide opportunities for tumor cells to evade immune surveillance. The negative correlation with naive B cells and resting mast cells is equally significant. The reduction in naive B cells and resting mast cells similarly reflects the formation of an immunosuppressive microenvironment. The decrease in naive B cells may reflect the inhibition of normal immune system function by the tumor microenvironment[58], while the reduction in resting mast cells may affect the initiation of innate immune responses[59]. Furthermore, the upregulation of Tregs is a particularly noteworthy finding. As key executors of immune suppression, Tregs enrichment in the tumor microenvironment is often associated with immune evasion and poor prognosis[60]. Although an increase in activated dendritic cells may represent the immune system’s attempt to initiate anti-tumor responses, such efforts may be effectively suppressed in a Tregs-dominant microenvironment[61]. Tumor microenvironment scoring analysis using the Estimation of STromal and Immune cells in MalignAnt neoplasm for ESTIMATE algorithm further confirmed the immune regulatory role of FOLR3. We found statistically significant differences in immune scores associated with FOLR3 expression, directly demonstrating the association between FOLR3 expression levels and the overall tumor immune microenvironment status.

A nomogram-based prognostic model constructed on the four key genes FOLR3, AURKA, POLQ, and CDKN2A demonstrated excellent clinical application potential. The model not only exhibited favorable calibration in the training set, but calibration curve validation showed good agreement between predicted and actual probabilities. The DCA further confirmed the clinical benefit value of the nomogram model. DCA, by comparing the net benefit of different decision strategies, provides clinicians with a quantitative decision support tool[62]. The model provides each patient with an individualized risk score, enabling clinicians to rapidly formulate appropriate management strategies based on the expression levels of the four genes. Furthermore, the consistency of all gene biomarkers functioning as risk factors significantly reduces the complexity of clinical application, facilitating clinician understanding and utilization. However, clinical application of the model still faces certain challenges. First, our study was primarily based on data from Western populations (TCGA predominantly comprises European and American populations), and the applicability in different ethnic and regional populations requires further validation. Second, gene expression may be influenced by multiple factors, including sample collection time, storage conditions, and detection methods, which may affect the stability and reproducibility of the model.

This study has the following limitations: First, the study was primarily based on retrospective bioinformatics analysis using TCGA and GEO databases, lacking detailed clinical follow-up data and protein-level validation. mRNA expression does not completely correlate with protein expression, particularly for membrane receptor proteins like FOLR3, where protein expression levels and cell surface localization may be more critical for functional manifestation. Second, samples were predominantly derived from European and American populations, and applicability in Asian populations such as China requires further validation, which has important implications for clinical translation. Third, functional mechanistic studies lack experimental validation. The predicted results of ceRNA regulatory networks and immune infiltration patterns require verification through cell experiments, animal models, dual-luciferase reporter assays, and RNA immunoprecipitation to confirm the authenticity of inter-gene regulatory relationships and elucidate the precise molecular mechanisms by which FOLR3 affects EC progression. Finally, immune infiltration analysis is based on algorithmic inference and may differ from actual immunohistochemical or flow cytometry detection results, particularly requiring cautious evaluation of cell types with unclear functional states.

## Author Contributions

Qin Tiansheng conceived, designed and supervised the study. Xu Xianyun performed the key bioinformatics analyses, interpreted the main findings, and drafted the manuscript. Wang Jiaxi collected and processed the data, and contributed to drafting the manuscript. Zhang Hongyu and Wang Yunyun assisted with bioinformatics analyses and results interpretation. Gao Xuelin validated the findings. ZZ and SL performed the molecular docking experiments. Qin Tiansheng and Xin Wenhu provided critical feedback and helped shape the research. Gao Xuelin revised the manuscript. Xu Xianyun acquired the funding. All authors discussed the results and approved the final manuscript.

## Data Availability

The complete dataset analyzed in this research is documented herein. Source data were extracted from publicly accessible genomic repositories (TCGA and GEO database). Derived datasets and computational outputs generated during the analytical workflow are obtainable from the corresponding author upon academic inquiry.

## Acknowledgments

We would like to express our sincere gratitude to all the participants for their contributions to this study.

