## Supplemental Data 1 for "Research on the Diagnostic Value and Immune Microenvironment Regulatory Mechanism of FOLR3 Gene in Endometrial Cancer Based on Multi-omics Data Algorithms"

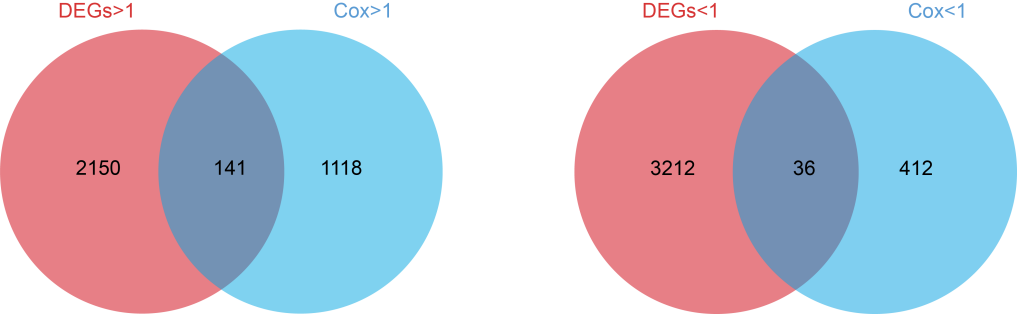

### **Supplementary Figure 1 Candidate gene screening**

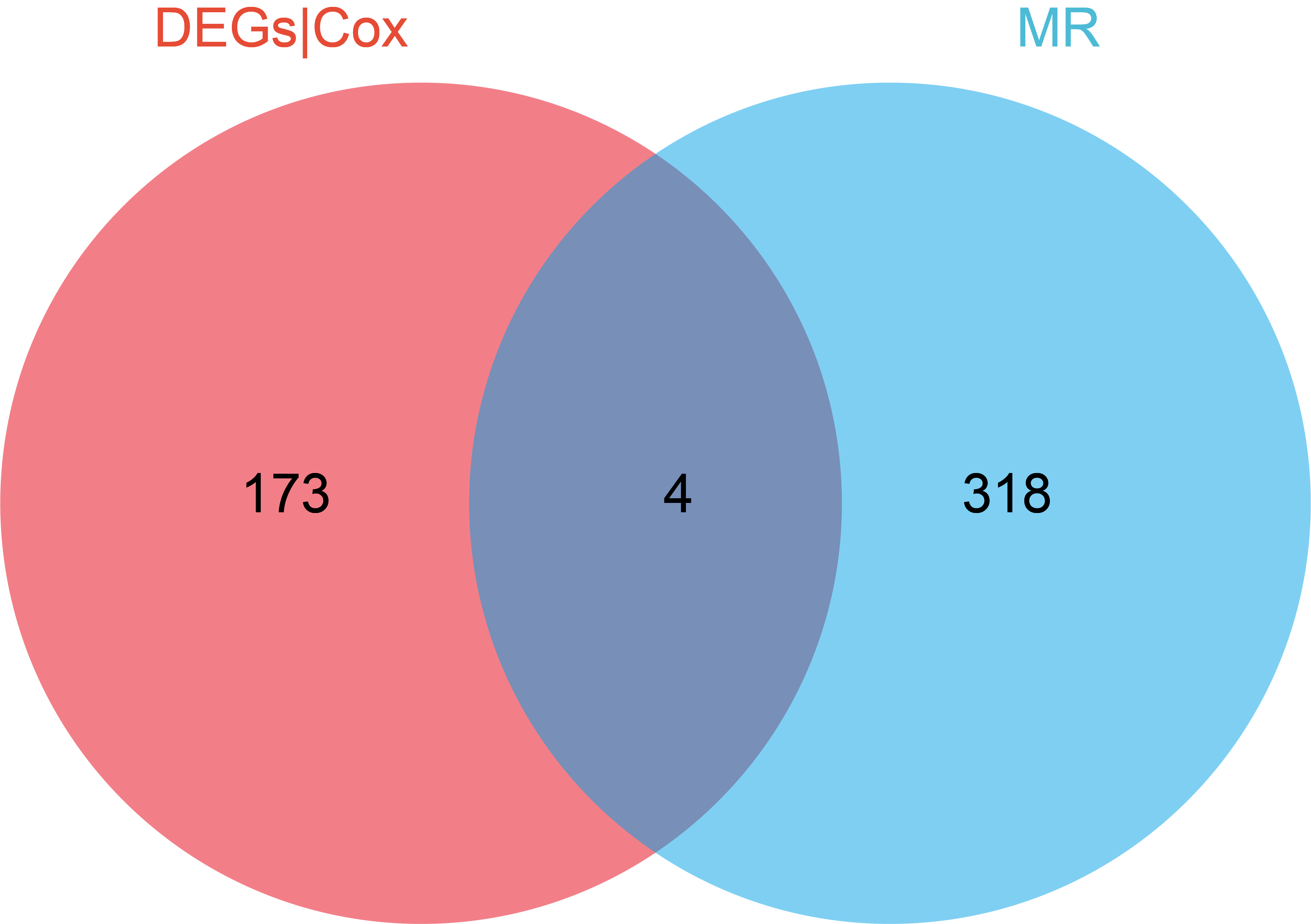

**Supplementary Figure 2 Intersecting genes screening**

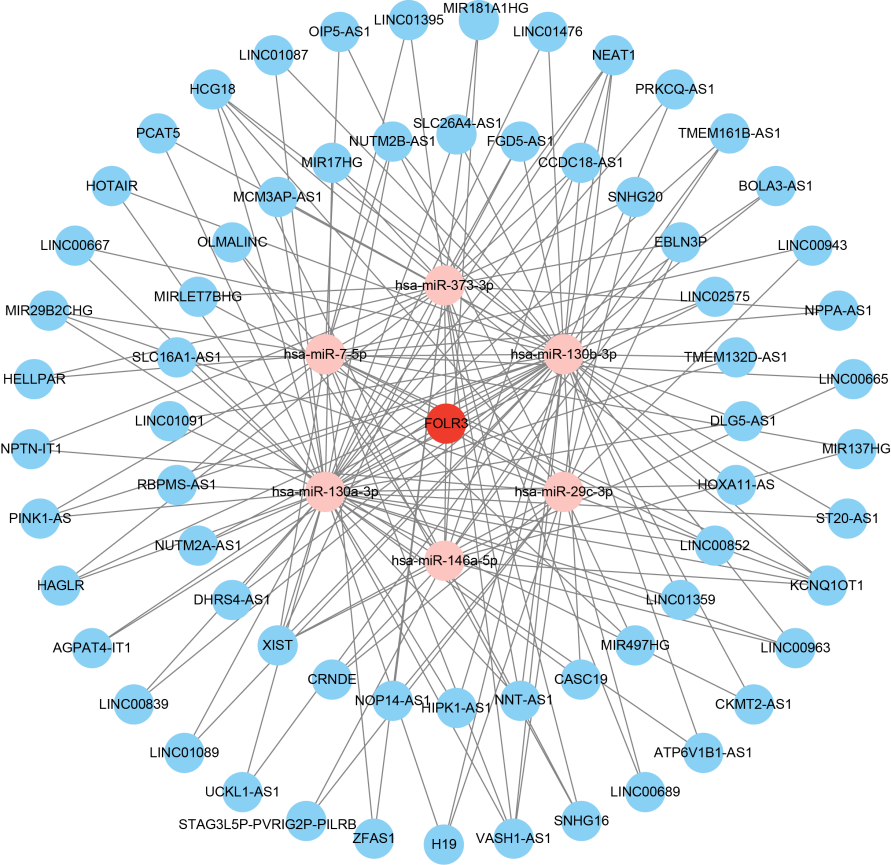

**Supplementary Figure 3 ceRNA Regulatory Network** FOLR3 gene-related ceRNA regulatory network. Red nodes represent FOLR3, pink nodes represent miRNA, and blue nodes represent lncRNA.
